# Clinical variables associated with immune checkpoint inhibitor outcomes in patients with metastatic urothelial carcinoma

**DOI:** 10.1101/2023.10.03.23296504

**Authors:** Soumaya Labidi, Nicholas Meti, Reeta Barua, Mengqi Li, Jamila Riromar, Di Maria Jiang, Nazanin Fallah-Rad, Srikala S. Sridhar, Sonia V. del Rincon, Rossanna C. Pezo, Cristiano Ferrario, Susanna Cheng, Adrian G. Sacher, April A. N. Rose

## Abstract

**Background:** Anti-PD-1/L1 immune checkpoint inhibitors (ICI) are indicated for metastatic urothelial cancer (mUC), however, only a minority of patients will derive therapeutic benefit. Strong predictive and prognostic factors are lacking. We investigated if clinical variables were associated with ICI outcomes in mUC.

**Methods:** We performed a multi-center retrospective cohort study of patients with mUC who received anti-PD-1/L1 ICI for metastatic disease between 2016-2021 at 3 Canadian cancer centres. Clinical characteristics, including demographics, BMI, metastatic sites, neutrophil-to-lymphocyte ratio (NLR), objective response, and survival were abstracted from chart review. ICI treatment response was determined by investigator assessment of clinical and radiologic parameters. Fisher’s exact test was used to assess differences in response rates between groups. Log rank and Cox regression models were used to assess overall survival (OS).

**Results:** We identified 135 patients with mUC who received anti-PD1/L1 ICI. A BMI ≥ 25 was significantly correlated to a higher overall response rate (ORR) to ICI (45.4% vs 16.3%, P = 0.020). After a median follow-up of 14.5 months, patients with BMI ≥ 30 experienced significant longer median OS 24.8 months vs. 14.4 months for 25 ≤ BMI < 30 and 8.5 months for BMI < 25 (P = 0.012). The ORR was significantly less in the presence of bone metastasis 16% vs 41% P = 0.006, and liver metastasis 16% vs 39% P = 0.013. Conversely, the presence of metastatic lymph nodes was significantly correlated with higher ORR 40% vs 20% P = 0.032. The median OS for patients with bone metastasis was 7.3 months vs 18 months in the absence of bone metastasis (P < 0.001). Patients with liver metastasis had a median OS of 8.6 months compared to 15 months for those without liver metastasis (P = 0.006). For lung metastasis, median OS was 8.7 months compared to 17.3 months (P = 0.004). No statistical difference was shown in OS for lymph nodes metastasis, with a median of 13.5 months vs 12.7 months (P = 0.175). Patients with NLR ≥ 4 had a significant worse OS (8.2 months vs 17.7 months P = 0.0001). In multivariate analysis, BMI ≥ 30, bone metastasis and NLR ≥ 4 were independent prognosis factor for OS.

**Conclusions:** Our data identified BMI and bone metastasis as novel, independent, clinical biomarkers that were strongly and independently associated with ICI response and survival in mUC. External validation of these data in a larger study and investigations into the mechanisms behind these findings are warranted.

## Introduction

Bladder cancer represents the 10^th^ most common cancer in the world, with approximately 550000 new cases annually, and accounts for 2.1% of all cancer deaths according to GLOBOCAN 2018 [1]. There is a male predominance, and smoking is the most common risk factor [1, 2]. Urothelial carcinomas arise from the urothelium of the bladder in most cases but can also develop from the upper urinary tract (renal pelvis and ureters). Tumors invading the detrusor muscle, ie., muscle invasive tumors, account with upfront metastatic disease for 25% of the cases [3]. Despite multimodality management for non-metastatic muscle invasive bladder cancer, 50% of these patients will relapse, with distant metastasis in most of the cases [4, 5]. Platinum-based chemotherapy remains the standard first line treatment for metastatic disease [4, 5]. Patients who respond or have stable disease following chemotherapy are eligible for subsequent maintenance therapy with the anti-PD-L1 immune checkpoint inhibitor, Avelumab [6], while those who fail to respond to chemotherapy subsequently can receive the anti-PD-1 immune checkpoint inhibitor, Pembrolizumab [7]. Other treatment options in the platinum-refractory setting include Enfortumab Vedotin, Erdafitinib in patients with susceptible FGFR alterations, and chemotherapy with taxanes (Paclitaxel, Docetaxel) or Vinflunine [8-11]. In the KEYNOTE-045 randomized phase 3 trial, Pembrolizumab showed a significantly longer overall survival of 10.3 months vs 7.4 months in patients with platinum refractory mUC compared to chemotherapy (Docetaxel, Paclitaxel, Vinflunine) (HR 0.73; 95%CI, 0.59 to 0.91 P = 0.002), with a lower rate of any grade adverse events (60.9% vs 90.2%) [7]. Long term results confirmed an overall survival benefit and a favorable safety profile [12]. The PD-L1 checkpoint inhibitor Atezolizumab was also associated with durable objective responses in a phase 2 trial, with favorable safety profile, [13] but failed to show survival benefit over chemotherapy in a phase 3 trial [14]. In phase 1/2 trials, Nivolumab, a PD-1 inhibitor, achieved objective responses alone or in combination with Ipilimumab, with a manageable adverse events profile [15, 16]. Despite responses seen with ICI in the platinum-refractory setting, not all patients will derive benefit, and there is a lack of valid prognostic and predictive biomarkers.

In this study, we aimed to evaluate if clinical prognostic markers, that have been defined for other cancer types, are relevant to mUC. In fact, several retrospective studies have addressed the relationship between obesity and outcomes in patients receiving ICI. In melanoma, non-small cell lung cancer (NSCLC) and renal cell cancer (RCC), obesity appears to be linked to better PFS and OS.[17] [18, 19]. And a recent systematic review of 18 retrospective studies across different cancer types, also found an association between high BMI and improved ICI outcomes; yet a strong positive correlation could not be concluded due to the heterogeneity of the studies [20]. As such, the relationship between BMI and immunotherapy outcomes in urothelial cancer remains poorly understood.

The tumor immune microenvironment of different metastatic locations may also affect the response to ICI [21, 22]. Retrospective studies reported differences in organ specific responses and organ specific overall survival with immunotherapy in several types of cancer [23-25].

Neutrophil-to-lymphocyte ratio (NLR) is an available marker of systemic response to inflammation, derived from the absolute counts of neutrophils and lymphocytes on a blood count [26]. It is a well-established poor prognostic factor in several cancers, independently of treatment type [26-28].

The aim of this retrospective analysis was to assess the association between high BMI, site of metastasis, NLR and outcome in terms of response and overall survival in a population of patients with mUC treated with ICI.

### Patients and methods

#### Patient population, characteristics, and outcome

We performed a multi-center retrospective cohort study analysis of mUC patients, from 3 Canadian cancer centers: Segal Cancer Center, Jewish General Hospital (JGH); Princess Margaret Cancer Center, University Health Network (PM-UHN) and Odette Cancer Center, Sunnybrook Health Sciences Center (SHSC). Patients with histologically proven metastatic urothelial cancer, who received at least one dose of anti-PD1/L1 ICI, between December 2016 and January 2021, were included regardless of gender, age, and performance status. ICI at any line of treatment for metastatic disease, alone or in combination with chemotherapy or another ICI, was allowed. Patients’ characteristics were abstracted from chart review. The following clinical characteristics were collected: age, gender, smoking status, comorbidities, primary tumor location and histology, type and line of ICI treatment, number and type of previous lines received, and sites of metastases. BMI at diagnosis, prior to ICI treatment and at progression was assessed. We selected 3 groups according to BMI at start of ICI: BMI < 25, 25 ≤ BMI < 30 and BMI ≥ 30. We collected the following outcomes criteria: ICI overall response rate (ORR), and overall survival (OS). ICI ORR was determined by investigator assessment of radiologic response as per RECIST criteria and was the sum of complete (CR) and partial response (PR). OS was calculated from the start of ICI treatment to death or last follow-up.

#### Ethics approval

Ethics approval for this study was obtained from the Clinical Trials Ontario (CTO Project ID: 2067) after review by the University Health Network Research Ethics Board on August 19, 2020, for PM-UHN and SHSC sites. Ethics approval for the JGH site was approved by the CIUSSS West Central Montreal REB (Project 2022-2888) on October 28, 2022.

#### Statistical analysis

Fisher’s exact test and chi square test were used to assess differences in response rates between the predefined groups. OS was assessed using Kaplan Meier. Log rank was used to compare groups and Cox regression models were used to perform univariable and multivariable analysis. The cut-off significant p value P < 0.05. Factors in the univariate analysis with a p value <0.05 and p value between [0.05-0.2] we included in the multivariate analysis.

## Results

### Patients’ characteristics

We identified 135 patients, who received at least one dose of ICI for mUC. The median age was 70 years (26-91). Most of the patients had a primary bladder cancer (92.5%). Most patients (n=84, 62%) received ICI as a second line or later treatment for mUC. The median follow-up period was 14.5 months. Patients’ characteristics for the entire cohort are shown in **Table 1**.

**Table 1.** Patients’ characteristics.

|  | N (%) |
| --- | --- |
| *Age* |  |
| Age ≤ 60 years | 28 (20.7%) |
| Age > 60 years | 107 (79.3%) |
| *Gender* |  |
| Male | 101 (74.8%) |
| Female | 34 (25.2%) |
| *Ever smoker* | 69 (51.9%) |
| *Primary* |  |
| Bladder | 125 (92.5%) |
| Upper tract | 10 (7.4%) |
| *Pathology* |  |
| Urothelial | 123 (91.1%) |
| Squamous | 7 (5.1%) |
| Other | 5 (3.7%) |
| *ECOG* |  |
| 0-1 | 112 (83%) |
| ≥ 2 | 15 (11.1%) |
| Missing data | 8 (5.9%) |
| *Metastatic sites* |  |
| LN | 90 (66.7%) |
| Lung | 48 (35.6%) |
| Bone | 43 (31.9%) |
| Liver | 37 (27.4%) |
| *BMI n=121* |  |
| < 25 | 55 (45.4%) |
| 25-30 | 43 (35.5%) |
| ≥ 30 | 23 (19%) |
| *ICI Line* |  |
| First | 51 (37.8%) |
| Second or later | 84 (62.2%) |

BMI data were available for 121 patients. Fifty five percent of the patients had a BMI ≥ 25. Median BMI was 20.9, 26.6 and 34.6 in groups BMI < 25, 25 ≤ BMI < 30 and BMI ≥ 30 respectively. At the time ICI treatment was initiated, metastatic sites were found as follows: lymph nodes 66.7% (n=90), lung 35.6% (n=48), bones 31.9% (n=43) and liver 27.4% (n=37).

### The relationship between BMI and ICI response and survival outcomes

We observed differences in ORR according to the BMI category of the patient. The ORR was 45.4% in the BMI ≥ 25 group, versus 16.3% in the BMI < 25 group (P = 0.020) (**Figure 1a**) In the BMI ≥ 25 and BMI < 25 groups, we observed a higher proportion of complete responses to ICI (8 CR vs 1, 22 PR vs 8 and 7 stable disease (SD) vs 10 (**Table 2**). Patients with BMI ≥ 30 experienced the longest median OS (24.8 months) compared with those with 25 ≤ BMI < 30 (14.4 months) and BMI < 25 (8.5 months) (P = 0.012). (**Figure 1b**). BMI≥ 30 remained an independent prognostic variable in multivariable analysis (HR = 0.40; 95% CI 0.17-0.96; P=0.040) (**Table 3**).

**Table 2.**
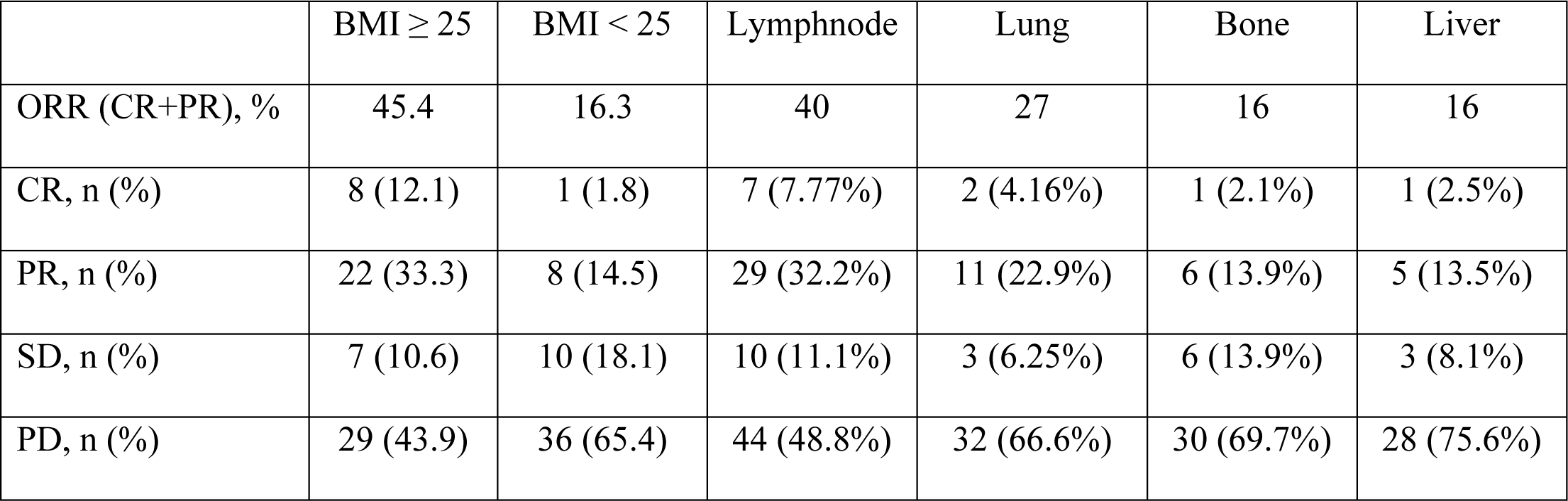
Responses according to clinical variables of BMI and metastatic sites.

| | BMI ≥ 25 | BMI < 25 | Lymphnode | Lung | Bone | Liver |
| --- | --- | --- | --- | --- | --- | --- |
| ORR (CR+PR), % | 45.4 | 16.3 | 40 | 27 | 16 | 16 |
| CR, n (%) | 8 (12.1) | 1 (1.8) | 7 (7.77%) | 2 (4.16%) | 1 (2.1%) | 1 (2.5%) |
| PR, n (%) | 22 (33.3) | 8 (14.5) | 29 (32.2%) | 11 (22.9%) | 6 (13.9%) | 5 (13.5%) |
| SD, n (%) | 7 (10.6) | 10 (18.1) | 10 (11.1%) | 3 (6.25%) | 6 (13.9%) | 3 (8.1%) |
| PD, n (%) | 29 (43.9) | 36 (65.4) | 44 (48.8%) | 32 (66.6%) | 30 (69.7%) | 28 (75.6%) |

**Table 3.** Uni- and Multivariable analysis for OS.

| OS | Univariable |  |  | Multivariable |  |  |
| --- | --- | --- | --- | --- | --- | --- |
|  | HR | 95% CI | p value | HR | 95% CI | p value |
| BMI ≥ 30 vs < 30 | 0.63 | 0.45-0.88 | 0.012 | 0.40 | 0.17-0.96 | 0.040 |
| Male vs Female | 0.95 | 0.58-1.57 | 0.867 |  |  |  |
| Age ≥ 60 vs < 60 | 1.88 | 0.65-5.45 | 0.235 | | | |
| ECOG ≥ 2 vs 0-1 | 1.68 | 0.86-3.29 | 0.125 | 2.21 | 1.02-4.78 | 0.042 |
| ICI Line 1 vs ≥ 2 | 2.39 | 1.43-3.97 | 0.001 | 1.80 | 1.31-2.48 | 0.000 |
| NLR < 4 vs ≥ 4 | 2.46 | 1.53-3.94 | 0.000 | 2.66 | 1.58-4.49 | 0.000 |
| Bone metastasis<br>Yes vs No | 2.38 | 1.52-3.73 | 0.000 | 1.98 | 1.17-3.35 | 0.010 |
| Lung metastasis<br>Yes vs No | 1.88 | 1.21-2.93 | 0.005 |  |  |  |
| Liver metastasis<br>Yes vs No | 1.92 | 1.2-3.09 | 0.006 |  |  |  |

**Figure 1.**
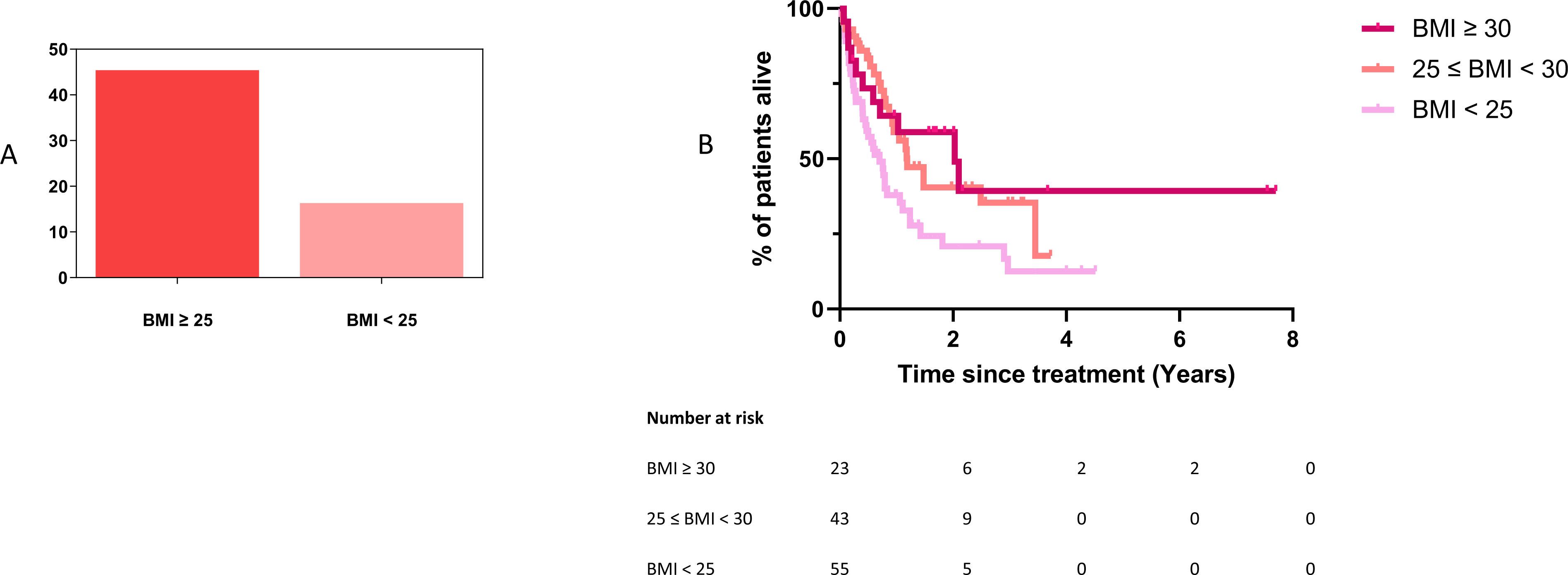
Overall response rate and overall survival according to BMI groups

### Metastatic sites’ response and survival outcomes

The overall response rate (ORR) for the entire cohort was 32.2%. The ORR was significantly lower in patients with bone metastasis 16% vs 41% (P = 0.006) (**Figure 2a**), or liver metastasis 16% vs 39% (P = 0.013) (**Figure 2b**). Conversely, the presence of metastatic disease involving the lymph nodes was significantly correlated with higher ORR 40% vs 20% (P = 0.032) (**Figure 2c**). The difference in ORR for lung metastasis was not statistically significant 27% vs 36% (P = 0.340) (**Figure 2d**). Detailed responses are listed in **Table 2**. The median OS was evaluated for the entire cohort, for each metastatic site and for each BMI group. In the entire cohort, median OS was 12.7 months. Bone, liver and lung metastasis correlated with significantly shorter survival. The median OS for patients with bone metastasis was 7.3 months vs 18 months in the absence of bone metastasis (P < 0.001) (**Figure 3a**). Patients with liver metastasis had a median OS of 8.6 months compared to 15 months (P = 0.006) (**Figure 3b**), and 8.7 months compared to 17.3 months for those with lung metastasis (P = 0.004) (**Figure 3c**). The presence of lymph node metastases was not significantly associated with OS: 13.5 months vs 12.7 months (P = 0.175) (**Figure 3d**). Although bone, lung, and liver metastasis were all associated with significantly worse survival in univariable analyses, bone metastasis was the only metastatic site that remained significantly associated with shorter overall survival in multi-variable analysis (HR = 1.98; 95%CI 1.17-3.35; P = 0.010) (**Table 3**).

**Figure 2.**
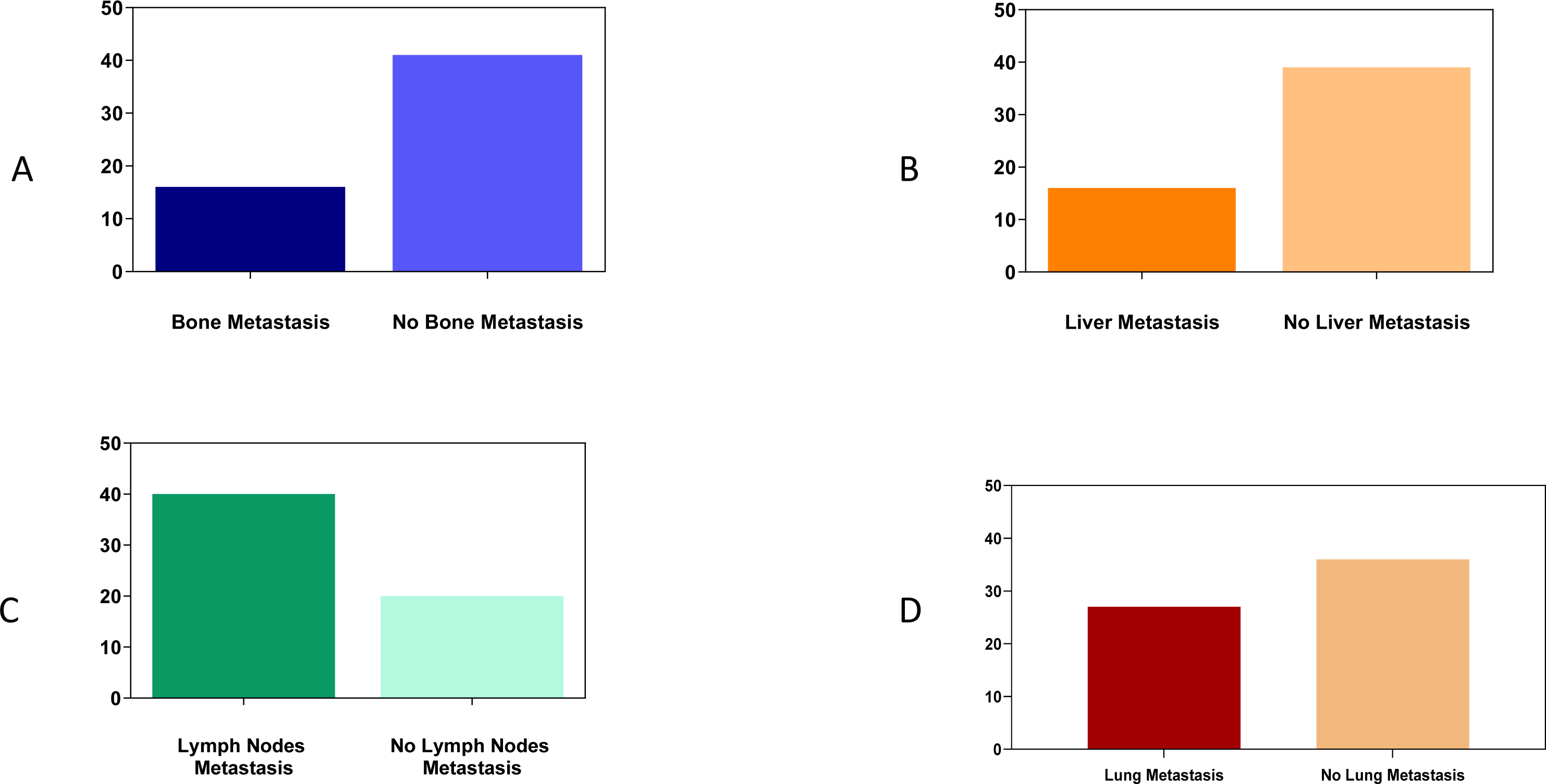
Overall response rate according to metastaticsites

**Figure 3.**
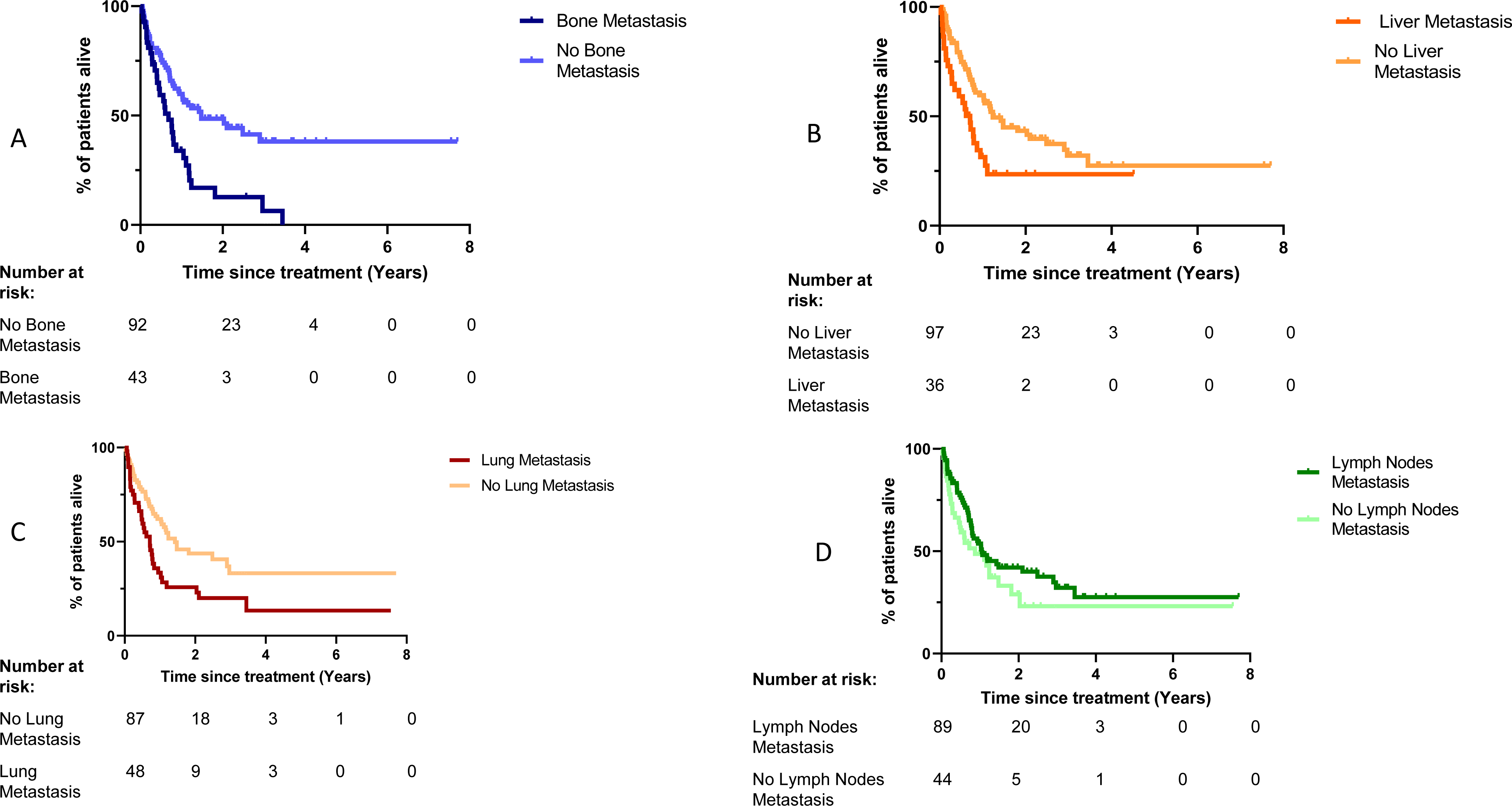
Overall survival according to metastaticsites

### NLR response and survival outcomes

Next, we evaluated the relationship between the neutrophil-to-lymphocyte ratio (NLR) and clinical outcomes with ICI in this cohort. The ORR was not statistically different for patients with NLR ≥ 4 compared to those with NLR < 4, respectively 24.5% and 36.9% (P = 0.141) (**Figure 4a**). However, NLR < 4 was correlated with better OS, with a median of 17.7 months versus 8.2 months for NLR ≥ 4 (P < 0.001) (**Figure 4b**). The survival benefit associated with a low NLR at the time of initiating ICI therapy was independent of other prognostic variable in multivariable analysis (**Table 3**). We assessed whether there was a relationship between NLR and BMI, but we did not observe any significant correlation (data not shown).

**Figure 4.**
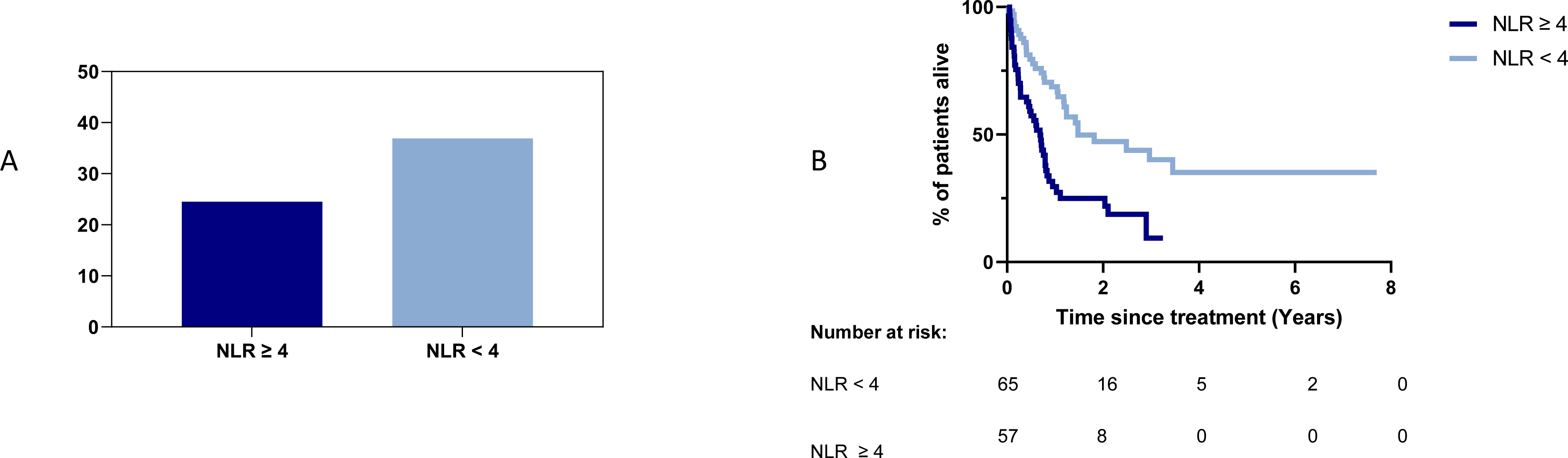
Overall response rate and overall survival according to NLR

## Discussion

ICI are approved for a wide-array of cancers based on tumor type (melanoma, NSCLC, urothelial cancer) and/or in a tumor type agnostic manner based on the presence of molecular biomarkers such as high tumor mutation burden (TMB) [29-31] and microsatellite instability [31-33]. However, in most cases, many patients who are eligible for immunotherapies based on tumor type or biomarker status will either not respond to treatment or will eventually become resistant to treatment. In this analysis, we identified several important clinical biomarkers that associate with better (high BMI) or worse (bone metastases, high NLR) outcomes with ICI in patients with mUC. These biomarkers are easily assessed without any additional costly molecular testing. Moreover, ICIs are expensive, potentially toxic, and in mUC only 30% of the patients benefit from ICIs. As such, we believe that these data may be clinically useful to guide treatment decisions.

According to the World Health Organization (WHO) obesity, classified as a body mass index (BMI) greater than 30 kg/m^2^, is a rising epidemic [34]. Our data showed strong correlation between elevated BMI, and improved outcome in terms of response as well as survival. To the best of our knowledge, the present study is the largest to report data of ICI outcomes correlation with BMI in mUC [20, 35, 36]. A favorable prognostic role of high BMI was also reported in RCC, NSCLC and melanoma [18, 35, 37-39]. A pooled post hoc analysis of individual participant data from 4 prospective trials with Atezolizumab in metastatic NSCLC showed significant difference in survival between normal weight, overweight and obese patients with improved OS for patients with obesity, and overweight compared to normal weight. [18] In a retrospective multicohort analysis of 1918 metastatic melanoma patients treated with chemotherapy, targeted therapies and ICI, McQuade et al studied association between BMI and outcome, as well as interactions between BMI and gender, and type of treatment. Obesity was associated with improved OS and PFS with a benefit restricted to patients treated with targeted therapies and ICI [17].

There is increasing evidence that excess body weight is a modifiable risk factor for several malignancies, including pancreas, kidney, colorectal, postmenopausal breast, ovarian, gallbladder and thyroid cancers [40]. However, obesity is not an established risk factor for bladder cancer, and the impact of obesity on cancer survival is less clear. Some studies have reported that obesity is associated with improved survival in some, but not all, patients with cancer, which is referred to the “obesity paradox” [41]. Numerous studies have shown that obesity can cause immune cell dysfunction [42-45], while others argue that the chronic low-grade inflammation underpinning obesity creates a pro-inflammatory state that synergizes with immunotherapy [46, 47].

Pre-clinical and translational research has provided insight into how immune checkpoint blockade can benefit obese patients. Obesity appears to influence T cell function and phenotype. For example, leptin-associated T cell dysfunction due to obesity has been observed across different species and tumor models, and was found to enhance the immune response to anti-PD-1 therapy [42]. Another mouse study reported that obesity caused a preferential increase and accumulation of senescent CD44^hi^CD62L^lo^CD4+ T cells that constitutively express PD-1 and CD153 [48]. Moreover, a study in a colon carcinoma mouse model showed that obesity impaired the infiltration, cytokine production and metabolic activity of tumor-infiltrating CD8 T cells, while anti-PD-1 restored CD8 T cells infiltration, proinflammatory cytokine production and metabolic activity, leading to complete tumor eradication and immune memory [49]. Obesity may also affect other immune cells in the TME, such as myeloid-derived suppressor cells (MDSCs), which are known to suppress T cell responses. MDSCs can suppress T cell activity through many mechanisms, one of which is expressing PD-L1 to induce T cell exhaustion [50, 51]. In a pre-clinical study, inhibiting PD-L1 on MDSCs decreased their ability to suppress T cell activity, suggesting that obesity could also accelerate tumor progression by promoting the expression of PD-L1 on MDSCs [52]. Finally, a study in a breast cancer model showed that although obesity accelerated tumor progression, anti-PD-1 treatment significantly reduced tumor burden by reshaping the local TME landscape [53].

The relationship between obesity and ICI is complex and depends on various of factors, including the types of cancer and ICI involved. However, BMI may be an imperfect marker and its evaluation based only on weight prior to ICI start can be limiting. Assessment of the weight loss or change over the time may be a better indicator of disease prognosis. As BMI cannot distinguish body fat from muscle, it may not be the best tool to assess obesity. Other measurement methods could be used for better assessment such as dual energy x-ray absorptiometry or fat referenced quantitative MRI [54].

Further studies are needed to understand the molecular mechanisms underlying the interaction between obesity and cancer immunity, and to identify potential targets for effective interventions.

Bone metastasis are a validated negative prognostic factor in mUC treated with platinum based chemotherapy [55, 56]. In our study, poor response and survival outcome of bone and liver metastasis was significant, while lymph nodes metastasis correlated with higher ORR. These results are consistent with published data in the literature for patients treated with ICI [57-59]. Makrakis et al. showed lower response rates and shorter OS for bone and liver metastasis on retrospective data from 917 mUC treated with ICI as 1^st^ or ≥ 2^nd^ line, but higher ORR for lymph node confined metastasis [57]. A retrospective multicentric Japanese study reported shorter ORR for bone metastasis in a cohort of patients treated with Pembrolizumab for mUC [58].Similar results have also been reported from prospective trials [60, 61]. In the IMvigor 210 phase 2 trial, the ORR on Atezolizumab was 32% for lymph nodes confined metastasis, and only 8% for liver metastasis, data for bone metastasis were not reported [60]. Higher ORR for lymph nodes metastasis (47%) compared to liver metastasis (23%) was also reported in the Keynote 052 trial [61]. Prognostic models for mUC patients treated with ICI identified liver or visceral metastases as poor prognostic factors [62, 63]. However, most prospective trials of ICI in mUC do not use presence/absence of bone metastases as a stratification factor, despite the fact that bone metastases are associated with poor response to chemotherapy, and impact quality of life [55, 56, 64]. Owari et al validated a specific prognostic scoring system, B-FOM, to predict survival for patients with bone metastasis from different genito-urinary cancers, based on 5 prognostic factors: primary tumor (prostate, renal or urothelial cancers), poor performance status, visceral metastasis, high Glasgow-prognostic score and elevated neutrophil-to-lymphocyte ratio [65]. This prediction tool may be helpful to individualize optimal treatment strategy for the patients. A better understanding of the bone microenvironment is crucial to help the development of novel therapeutic strategies and improve outcomes [66].

The distant organ microenvironment, also known as metastatic microenvironment (MME), plays an important role in site-specific metastasis [67]. For instance, macrophages, which are abundant immune cells in the lungs, can bind to cancer cells via receptor VCAM-1 transmit, signaling a chain of events leading to lung-specific metastasis in breast cancer [68]. It has been demonstrated that myeloid cells can remodel the pre-metastatic lung from an immune protective state to a state favoring tumor progression, thereby promoting lung metastasis [69]. Furthermore, lung stromal cells can also promote tumor colonization and metastasis by secreting periostin [70]. In the liver, there is a significant presence of circulating and resident NK cells, which serve as the primary effectors of liver immune function. This NK cell maintained the breast cancer cells dormant in the liver by secreting IFNγ, while sustaining NK cell abundance with IL15-based immunotherapy succeeded to prevent liver metastasis and prolong the survival in preclinical models [71]. Conversely, bone forms an immunosuppressive environment mainly due to immature NK cells and a small number of cytotoxic T cells, and a large number of myeloid progenitors and Treg [72]. In a human bladder tumor xenografts model, researchers observed a high infiltration of bone marrow-derived host CD11b myeloid cells. They further demonstrated that enhanced tumor-associated in the TME promote an immunosuppressive pro-tumoral myeloid phenotype [73]. These observations demonstrate how the MME can influence the metastatic potential and outcome of different types of cancer cells in different organs, suggesting that MME might also potentiate site-specific metastasis in bladder cancer. Taken together, all these studies suggest the importance of understanding the metastatic potential and outcome of different types of cancer cells in different organs, through which we can further develop more effective therapeutic measures against bladder cancer.

NLR is a well-established poor prognostic factor in several cancers, independently of treatment type [26-28]. It could be considered as a surrogate marker of chronic inflammation and evasion of immune surveillance [74]. In our study, high NLR was associated with shorter median OS. A recent study by Valero et al showed poor response and survival outcomes, for 1917 patients with high NLR at diagnosis, treated with ICI for multiple cancer types [74]. Similar results were reported for melanoma and NSCLC [27, 75]. Banna et al explored the prognostic and predictive role of NLR and LDH in metastatic urothelial cancers treated with ICI, and showed significant correlation with PFS and OS for high NLR [76]. For patients treated with ICI for mUC, a high NLR prior to first line chemotherapy and second line pembrolizumab was associated with worse survival outcomes [77]. NLR may represent an accessible low-cost predictive biomarker.

The present study has some strengths and limitations. It presents a relatively large multi-center cohort of patients with significant prognostic factors. On the other hand, this is a retrospective study, with heterogenous population, and an investigator-based response assessment on clinical and radiological chart review. However, these observations made in this analysis are hypothesis generating. The evaluation of obesity and its relationship with ICI response warrants future prospective studies that incorporate better established measures of obesity and metabolic syndrome. Moreover, little is known about how bladder cancer cells interact with the liver and bone tumor immune microenvironments and this warrants further investigation in preclinical models to develop optimal immunotherapies for patients with bone and liver mUC.

## Conclusion

Our study identified elevated BMI, NLR and presence of bone metastases as potential biomarkers for ICI response and survival in mUC. Obesity was associated with improved survival and response rate, whereas bone metastases and high NLR are associated with lack of response and shorter survival. Prospective validation of these data is warranted, especially in the evolving landscape of therapeutic options for mUC with novel agents and combinations with ICI [78].

## Data Availability

All data produced in the present study are available upon reasonable request to the authors

